# Neural sensitivity in cochlear implantees determined by electrically-evoked compound action potentials (ECAP) and focused perceptual thresholds

**DOI:** 10.1101/2025.10.30.25338964

**Authors:** Dietmar M. Wohlbauer, Julie G. Arenberg

## Abstract

**Purpose:** The sensitivity of auditory neurons in response to electric stimulation in cochlear implant (CI) listeners may reflect neuronal health, which is a major contributor to variability in performance outcomes among CI listeners. In the current study, we explore the interplay of three outcome measures of neural sensitivity, perceptual focused thresholds, objective electrically-evoked compound action potentials (ECAP), and the Failure Index (FI). We further explore the influence of CI experience and how the measures may contribute to speech perception performance.

**Methods:** We examined focused perceptual threshold measures, ECAP stimulation levels and N_1_P_2_ peak response amplitudes, and ECAP input-to-output relationships in 29 adult CI recipients (14 females, 11 males, four who were bilaterally implanted). Pearson correlation analysis was performed to investigate subject specific relationships across CI electrodes, and linear mixed-effects models (LMM) were used to identify links between the outcome measures accounting for individual variation.

**Results:** Individual outcomes revealed large within and across subject variability for focused thresholds, ECAP peak amplitudes, and FI. The LMMs showed, that low focused threshold measurements correspond to large ECAP peak amplitudes, while high ECAP stimulation levels reflect large ECAP peak amplitudes. Furthermore, clinical speech perception seems to be influenced by the relationship of focused thresholds and FI, with lower performance for stronger associations.

**Conclusion:** Our findings suggest that the combination of objective ECAP response measures and perceptual measures could be a robust estimate of neural health and may act as an early estimate of the speech performance abilities in CI listeners.

## 1 Introduction

Cochlear implants (CI) are neural prostheses widely used to restore hearing of individuals with severe to profound hearing loss. CIs enable sound perception by capturing acoustic signals via one or more microphones, processing the available signal to emphasize specific signal components, and distributing the extracted information to a CI electrode array. The electrode array is inserted in the cochlear of the inner ear in close proximity to the auditory nerve to electrically stimulate groups of auditory neurons. These neurons generate electrical patterns via series of action potentials upon activation, which are further transmitted to the brain for higher level processing. Even though that electric signal transmission via CIs is highly successful with more than one million implantees worldwide [1], performance outcomes such as speech perception, are below those of normal hearing individuals and show large variations across CI listeners.

Some factors that lead to the observed variability in performance outcomes are rooted in the nature of the CI implantation, dependent on the anatomy of the human temporal bone and cochlea and the CI technology itself. CI surgery is a highly individualized procedure, and differences in electrode location and insertion depth across CI candidates (e.g., [2–4]), or the applied signal processing that converts acoustic to electric signals (e.g., [5–7]) are just some influential factors in this context.

A key factor that limits the hearing performance in CI listeners is the auditory neural health, that is the decrease in the density or responsiveness of auditory neurons along the length of the cochlea and corresponding degeneration of the auditory nerve, following sensorineural hearing loss (e.g., [8–10]). This neural degeneration can vary along the cochlea and electrically stimulating a dense or “good” versus sparse or “poor” neural area with a CI, leads to differences in the corresponding evoked neural activity and the effectively transmitted signal information. Hence, perceptual performance measures of CI listeners depend on the sensitivity of the auditory neural region to electric stimulation.

Several studies have investigated the anatomical state of the auditory neurons in animal models [11, 12], and measures of evoked responses showed the functional relation between surviving neurons and response amplitudes (e.g., [13, 14]). With CIs, this neural status might be estimated directly through the electrode array by recording electrically-evoked compound action potentials (ECAP), an objective assessment reflecting the synchronous response of the auditory neurons to electrically applied stimuli [15–19]. ECAPs can be recoded with a forward-masking stimulus complex of different masker and probe electrode combinations [20–23], and have been used more recently as a measure of neural sensitivity via the quality of the electrode-neuron interface (ENI), that is the interface between CI electrode and auditory neurons, hence, as a potential marker for auditory neural health.

A number of studies explored neural sensitivity in the animal model via the temporal aspects of electric stimulation (e.g., [15, 16]). Briefly, CIs use biphasic stimulation pulses to elicit a hearing sensation. Individual pulses often comprise an interphase gap between the first and second phase and varying the duration of this gap influences the excitability of the corresponding neural population. This phenomenon is called the interphase gap effect (IPG). Healthy neural populations show larger IPG effects than degenerated populations, especially when comparing ECAP amplitude growth with increasing stimulation intensity [16].

Another approach to investigate neural sensitivity in animal models is to introduce a microlesion in the spiral ganglion neurons, as was done by Konerding and colleagues (2025) [19]. They demonstrated that neural sensitivity and a neural lesion could be estimated via the place-specific relationship of the input stimulation level to output peak neural response amplitude, denoted as the Failure Index (FI). Similarly, the FI can potentially capture the neural responsiveness and variability across the electrode array in human CI listeners. Hence, low FIs would reflect good neural sensitivity and ENI, whereas high FIs would correspond to a poor ENI and less sensitive regions.

The assessment of the integrity of the auditory neurons in animal models has been shown to be most important to gain direct insights into the effects of the neural sensitivity in humans (e.g., [24]). The interpretation of auditory health in human subjects, however, is more challenging than in animal models because the spiral ganglion neurons cannot be imaged in detail. Nevertheless, the translation from neural activity to the corresponding survival in humans via physiological ECAP recordings has shown some promise (e.g., [25–27]). McKay and Henshall [25] showed an increased neural sensitivity with longer IPGs. Later, He et al. [26] demonstrated, that increasing the IPG improves neural responsiveness for children with cochlear nerve deficiency, however, IPG effects might depend more strongly on the health status of the auditory nerve than the neurons itself. Therefore, the IPG might not solely reflect the ENI but a combination of peripheral and more central processes. Other studies investigated the ECAP spread of excitation (SOE) as a possible marker for evaluating the auditory neural survival in CI listeners ([4, 17, 18, 28]. SOEs represent the activation of a neural population along the ENI to a place-specific probe stimulus, resulting in a spatial representation of neural activity. These studies showed large variability in neural activity across electrodes and subjects, suggesting that accurate information about the ENI is needed, to understand the main mechanisms in CI stimulation and to improve performance outcomes. As mentioned before, Konerding et al. [19] proposed a neural health index based on the relationship between input stimulation current level to output neural response amplitude of a masker-probe pair of ECAP measurements. In that study, the FI was measured at the response saturation level of the ECAP AGF. While saturation can be reached in anesthetized animals, this level would likely result in stimuli exceeding the maximum acceptable loudness in awake humans. Nevertheless, the standard deviations (SD) of the FI were robust at lower levels along the AGF down to 70%, a possible indicator, that FIs extracted at lower stimulation levels in humans might still reflect accurate ratios. Recently, Garcia et al. [29] presented a comparison between the FIs and panoramic-ECAP (PECAP, [18]) neural health measures in a population of human listeners. The study found, that both FIs and PECAP outcomes identified regions along the CI array with good and poor neural sensitivity, potentially reflecting neural health. In a comparison with computerized tomography (CT) scans and calculations of the electrode-modiolus distance [30], the study indicated, however, that the FI might be dependent on the distance of the electrode array to the auditory nerve, while the PECAP estimate does not show this dependence. Hence, a robust estimation of neural health in humans might need an extension to other measures of auditory performance, to clarify the identified discrepancies.

Beside physiological ECAP recordings, the ENI was also investigated via various perceptual measures. Differences in neural sensitivity were shown in the temporal domain via multipulse integration (MPI, e.g., [31, 32]), as well as in the spectral domain via psychophysical tuning curves (PTC, e.g., [33, 34]) or perceptual thresholds (e.g., [35–39]). Specifically, Bierer et al. [38] suggested to use focused perceptual thresholds as a psychophysical method to assess the neural sensitivity to focused electrical stimuli. The measurement uses a steered quadrupolar electrode configuration, that is a combination of four electrodes, where the stimulation current of the two central electrodes is stepwise steered from one to the other electrode, while the two flanking electrodes are phase inverted with a constant current, a method to focus the electric field to a narrower neural area. By measuring the detection threshold of focused stimuli along the CI electrode array, it was shown, that focused thresholds capture larger threshold variations across electrodes, a possible indicator for the neural sensitivity of the corresponding cochlear region [4, 34, 37]. Recently, Peng et al. [39] investigated the difference between bipolar and monopolar perceptual threshold measurements along the CI electrode array as a possible neural health metric, with large threshold differences indicating regions with poor neural health and vice versa. In a comparison with PECAP neural health estimates the study found correlates on individual and group levels between the two measures, suggesting that perceptual thresholds and PECAP share similar aspects as neural measures.

Compared to animal models, which are quite limited to an investigation of neural health based on anatomical and physiological features, human CI performance can be assessed subjectively, i.e. speech perception. Several of the above-mentioned methods highlighted the impact of neural health measures on perceptual performance outcomes. Specifically, the ECAP IPG effects, SOE and channel interaction between electrodes revealed its sensitivity to perceptual performance outcomes, such as pitch ranking, speech perception, or spectral resolution (e.g., [28, 40, 41]). Also, some perceptual measures of neural sensitivity to electric stimulation, such as focused perceptual thresholds, correlated with speech performance or spectral resolution outcomes (e.g., [39, 42]), while other measures did not show a relation between neural measures and speech performance (e.g., [43]).

In the current study we evaluate, if three outcome measures, perceptual focused thresholds, objective ECAP recordings of the auditory nerve, and from the ECAP derived FIs, (A) address similar aspects of neural sensitivity, and (B) can be combined to get a more robust estimate of the neural sensitivity across CI electrodes. Furthermore, we test if the relationship between focused thresholds and the FI relates to clinical speech performance scores and duration of CI listening experience. Our findings intend to create a foundation for potential new methods that can be used to investigate the ENI in a clinical setting, with the aim to improve signal transmission through either CI programming including channel deactivation, or novel CI stimulation methods, and the long-term goal to improve CI performance and listening satisfaction.

## 2 Methods

### 2.1 Participants

Twentynine adult CI recipients (14 females, 11 males, four who were bilaterally implanted) recruited from the Massachusetts Eye and Ear, Department of Audiology, and the preexisting cochlear implant database of the laboratory, participated in the current study. Participants were speakers of American English speakers and were implanted with Advanced Bionics (AB) the HiRes90K device and used the HiRes Optima-S as their everyday coding strategy. Clinical consonant-nucleus-consonant (CNC) performance ranged from 12% to 88% correct words with the average at 66.2%. Participants had more than 6 months experience with their implant, with an average of 7.5 years. Clinical speech scores, demographics, etiologies, and the mean focused thresholds are shown in Table 1.

**Table 1:** Study participant demographics, sex, age at testing, CI experience at testing, etiology, CI array type, consonant-nucleus-consonant (CNC) score, ECAP stimulation phase duration. Participants are sorted based on average focused perceptual thresholds across CI electrodes, to align with our previous studies.

| # | Subject ID* | Sex | Ear | Age<br>(range, years) | CI experience<br>(years) | Etiology | CI type | CNC<br>(% correct) | Average focused<br>threshold (dB re 1 μA) |
| --- | --- | --- | --- | --- | --- | --- | --- | --- | --- |
| 1 | A041 | F | R | 66-70 | 8.2 | Genetic | Mid-scala | 70 | 55.82 |
| 2 | A028 | F | R | 66-70 | 5.8 | Unknown | SlimJ | 82 | 53.56 |
| 3 | A043 | M | L | 71-75 | 2.8 | Unknown | Mid-scala | 84 | 52.62 |
| 4 | A039 | F | L | 51-55 | 3.4 | Ototoxic | Mid-scala | 68 | 51.62 |
| 5 | A023 | M | L | 66-70 | 2.0 | Genetic | Mid-scala | 32 | 50.46 |
| 6 | A005 | F | L | 71-75 | 11.8 | Hereditary | Helix | 80 | 49.43 |
| 7 | A042 | M | R | 21-25 | 2.6 | Unknown | Mid-scala | 46 | 49.39 |
| 8 | A040 | F | L | 26-30 | 4.9 | Ototoxic | SlimJ | 36 | 49.38 |
| 9 | A022 | M | L | 56-60 | 20.3 | Genetic | 1J | 80 | 48.53 |
| 10 | A019 | F | R | 76-80 | 5.5 | Autoimmune | Mid-scala | 26 | 48.37 |
| 11 | A022 | M | R | 56-60 | 21.2 | Genetic | 1J | 80 | 48.02 |
| 12 | A007 | M | R | 56-60 | 2.4 | Menière's | SlimJ | 12 | 47.82 |
| 13 | A029 | F | R | 66-70 | 1.4 | Unknown | Mid-scala | 84 | 47.69 |
| 14 | A020 | F | L | 61-65 | 10.1 | Hereditary | Helix | 82 | 47.59 |
| 15 | A024 | F | L | 76-80 | 6.7 | Menière's | Mid-scala | 62 | 47.52 |
| 16 | A002 | M | L | 61-65 | 19.4 | SSNHL** | Mid-scala | 92 | 47.27 |
| 17 | A023 | M | R | 66-70 | 7.4 | Genetic | Mid-scala | 68 | 45.41 |
| 18 | A002 | M | R | 61-65 | 17.1 | SSNHL** | Helix | 88 | 45.30 |
| 19 | A015 | F | L | 56-60 | 8.2 | Unknown | Mid-scala | 58 | 45.13 |
| 20 | A029 | F | L | 66-70 | 10.2 | Unknown | Mid-scala | 76 | 44.40 |
| 21 | A027 | M | L | 41-45 | 1.4 | Unknown | Mid-scala | 70 | 43.67 |
| 22 | A035 | M | R | 71-75 | 0.7 | Genetic | Mid-scala | 70 | 42.43 |
| 23 | A034 | F | R | 66-70 | 4.4 | Genetic | Mid-scala | 86 | 41.64 |
| 24 | A017 | F | L | 51-55 | 6.6 | Ototoxic | Mid-scala | 52 | 40.33 |
| 25 | A016 | M | L | 36-40 | 3.1 | Hereditary | SlimJ | 88 | 40.14 |
| 26 | A013 | M | L | 31-35 | 2.1 | Hereditary | Mid-scala | 64 | 39.74 |
| 27 | A011 | F | L | 56-60 | 16.9 | Viral | Helix |  | 39.50 |
| 28 | A008 | M | R | 26-30 | 4.1 | Unknown | Mid-scala | 62 | 38.30 |
| 29 | A014 | F | L | 66-70 | 8.1 | Menière's | Mid-scala | 56 | 38.10 |
| *Mean ± SD* | | | | 58.8 ± 15.3 | 7.5 ± 6.1 | | | 66.2 ± 20.5 | 46.18 ± 4.71 |

\* sorted with decreasing average focused threshold \*\* SSNHL (Sudden sensorineural hearing loss)

### 2.2 Focused Perceptual Thresholds

Focused perceptual threshold measurements were performed with the previously reported fast psychophysical sweep procedure [38]. The participant’s internal device was connected via a dedicated laboratory Advanced Bionics (AB) Naida Q90 research processor through the Clinical Processing Interface (CPI3, Advanced Bionics Corp.). Stimuli were be controlled through the Bionic Ear Data Collection System software (BEDCS, AB Corp., currently version 3.0) installed on a research desktop computer. The upper stimulation limits per CI electrode were defined behaviorally prior to the focused threshold measurement via the assessment of the “most comfortable loudness” (MCL) level for the focused stimuli for electrodes 2 to 15. By using AB’s clinical rating scale subjects were asked to rate stimuli loudness for stimuli presented at a steadily increased stimulation level until loudness rating 7, “loud but comfortable”. The final limit was then defined as rating of 6 or “most comfortable”, and the initial presentation level was set to 3 dB below the MCL level, to reduce the risk of too loud stimuli during the automatic procedure. The sweep procedure for the focused threshold measurement uses a steered quadrupolar (sQP) electrode configuration, with sigma (*σ*) controlling the return current for the two flanking electrodes and extracochlear ground, and a steering coefficient alpha (*α*), that controls steering between the apical and basal electrodes of the sQP configuration. For example, a value of *σ* = 0.9 returns 90% of the current to the flanking electrodes, and 10% to the extracochlear ground. A value of (*α*) = 0 delivers the full current defined by *σ* to the apical electrode, an *α* = 0.5 shares the current equally between apical and basal electrodes, and *α* = 1 delivers the full current to the basal electrode only. Cathodic leading biphasic pulses of 97 µs pulse width and 0 µs interphase gap per phase are presented as pulse trains of 200.4 ms with regular intervals of 997.9 pulses per second, and with increasing *α* in 0.1 steps from the most apical set to the most basal set of electrodes. The steady change of stimuli is triggered by the computer keyboard’s spacebar, which either decreases its stimulation level by pressing or increasing the level by releasing the spacebar. A full threshold measurement consists of averaging the results of two sets of forward sweeps (apex-to-base) and backward sweeps base-to-apex).

### 2.3 Electrically-evoked compound action potential

Electrically-evoked compound action potentials (ECAP) were recorded with the same AB hardware and software as used for the focused threshold recordings and by using the common forward-masking technique [18, 20, 22, 23]. In brief, matching masker-probe electrode pairs were defined for all clinically active electrodes and ECAP responses derived from the recordings of four stimulation sequences containing different combinations of masker and probe stimuli. The ECAP recording channel was located apically from the active probe channel with a constant offset of two electrodes, except when the masker-probe pair was located at the most apical electrodes, then the recording locations were moved to the basal side of the pair. Default ECAP stimulation parameters were used for all recordings, including pulse width (21.55-53.88 µs), masker-probe interval (600 µs), and stimulation rate (80 pulses per second). All recordings were repeated 50 times per masker-probe condition and averaged to reduce recording noise. Prior to the ECAP recording, the loudness growth of the ECAP stimulus sequences was obtained for all clinically active electrodes similarly as described for the focused threshold measurement, to determine the upper limit of stimulation level for the experimental task indicated by a rating of 7 or “loud but comfortable”, to prevent participant discomfort. A custom graphical user interface developed in MATLAB was used to perform the ECAP experiments. Stimulation parameters are sent via the user interface and BEDCS to the research processor to stimulate the implant sequentially with the prepared sequences. The resulting neural recordings are downloaded from the processor and stored in stimulation matrices, containing all individual ECAP frames, information then used to extract the ECAP responses. This process was repeated for all matching masker-probe pairs and N_1_P_2_ peak response amplitudes were extracted from the average ECAP measurement.

### 2.4 The Failure Index

Following the ECAP measurement, the Failure Index (FI) was calculated for all active electrodes based on Konerding et al. [19] (Eq. 1). To account for the variability between study participant, we calculated the FI ratio in dB-converted stimulation levels (dB re 1 µA) and ECAP peak response amplitudes (dB re 1 µV). The calculations used ECAP stimulation currents at “loud but comfortable” levels or rating of 7 on AB’s clinical loudness rating scale, indicated as LC in (Eq. 2). In contrast to Garcia et al. [29], who suggested to use the stimulation charge calculated from current levels (in µA) at MCL and stimulation phase (in µs) to account for larger measurement differences in humans than in the animal model, our calculations closely match the initially suggested approach.

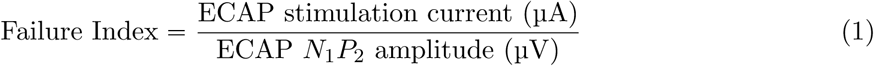

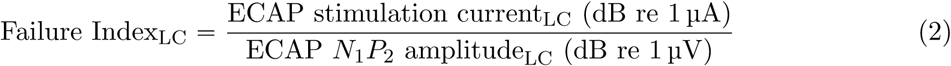

### 2.5 Statistical Analyses

All statistical analyses were performed in MATLAB 2023b. Statistical significance was defined as p ă .05. Focused perceptual threshold and ECAP parameters (stimulation current and N_1_P_2_ peak response amplitude) were first investigated on a subject-to-subject basis. Pearson correlation coefficients were computed between focused thresholds and ECAP peak amplitudes, between ECAP peak amplitudes and ECAP stimulation levels, and between focused thresholds and FI calculations. Individual linear regression analyses via MATLAB’s *fitlm* function were conducted beside partial correlations, to examine whether electrode location predicts focused thresholds and ECAP peak amplitudes. Based on these initial investigations, four linear mixed-effects models (LMM) were fitted via MATLAB’s *fitlme* function, to measure the relationship between (A) focused thresholds and ECAP peak amplitudes across the study population, (B) ECAP stimulation current level and ECAP peak amplitudes, (C) the relationship of focused thresholds on the ECAP input-to-output ratio (i.e., the Failure Index), and (D) the effects of clinically assessed speech (CNC scores) and CI experience on the relationship between focused threshold and FI. Between-participant random intercepts were included in all models. Co-linearity of CNC scores and CI experience was tested via Kendall’s *τ* . The final LMM equations are shown below (Eq. 3, 4, 5, 6). Marginal and conditional *R*^2^ to assess the variance of fixed effects alone and both fixed and random effects were calculated after [44].

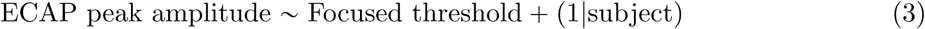

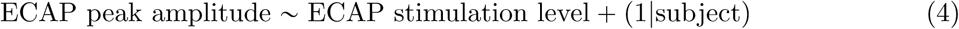

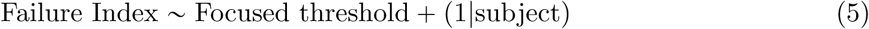

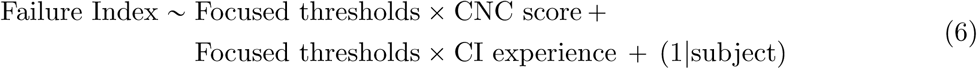

## 3 Results

To highlight the measurement variability across electrodes and subjects, we picked two example participants. Fig. 1 shows ECAP recordings resulting from the forward-masking procedure. Our recordings show that ECAP responses are highly variable across electrodes and participants. A007 / R shows largest ECAP responses in the middle of the array indicated by blue N_1_ troughs and yellow P_2_ peaks and a maximum peak amplitude of 378.7 µV, whereas A008 / R shows large basal responses with a maximum peak amplitude of 525.5 µV.

**Figure 1:**
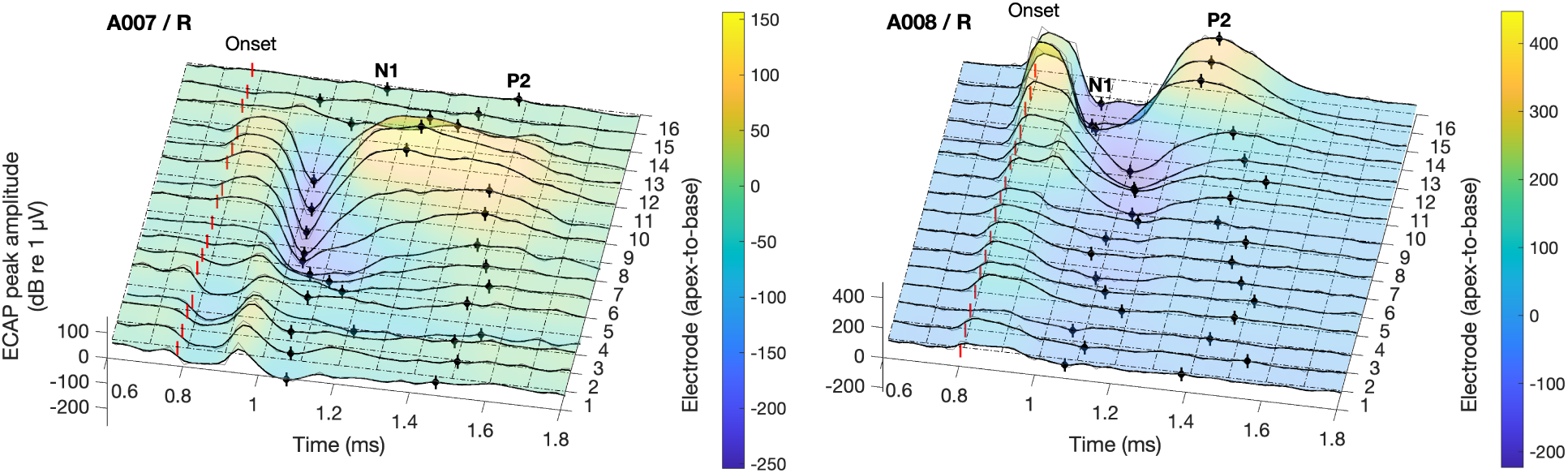
ECAP responses of two example participants, A007 / R (left panel) and A008 / R (right panel), All available electrodes from apical to basal on the y-axis, and assessment time frame between 0.6 and 1.8 ms on the x-axis, and the color map representing ECAP peak response amplitudes in µV to the right of each panel. ECAP probe onsets, N_1_ troughs and P_2_ peaks indicated per electrode with red and black markers per electrode and response.

For the same two example participants, outcomes for all investigated parameters are shown in Fig. 2 (top panels) with focused perceptual threshold profiles (dB re 1 µA, green filled circles), ECAP stimulation current levels (dB re 1 µA, blue open circles), and ECAP peak amplitudes (dB re 1 µV, blue filled circles). Pearson correlation outcomes are shown for focused thresholds versus ECAP peak amplitudes (*r/p*_A_), and for ECAP input stimulus current versus output amplitude (*r/p*_B_). Note that A007 / R (left panels) shows a significant inverse relationship for thresholds and ECAP peak amplitudes (*r* _A_ = –.71, *p*_A_ = .007), hence, low thresholds reflect high ECAP peak amplitudes and vice versa, and a significant positive relationship for ECAP input stimulus current versus output amplitude (*r* _B_ = .75, *p*_B_ = .002). In contrast, A008 / R (right panels) does not show clear relationships between the different variables, neither between focused thresholds and ECAP peak amplitudes, nor between ECAP input stimulus current and output amplitude.

**Figure 2:**
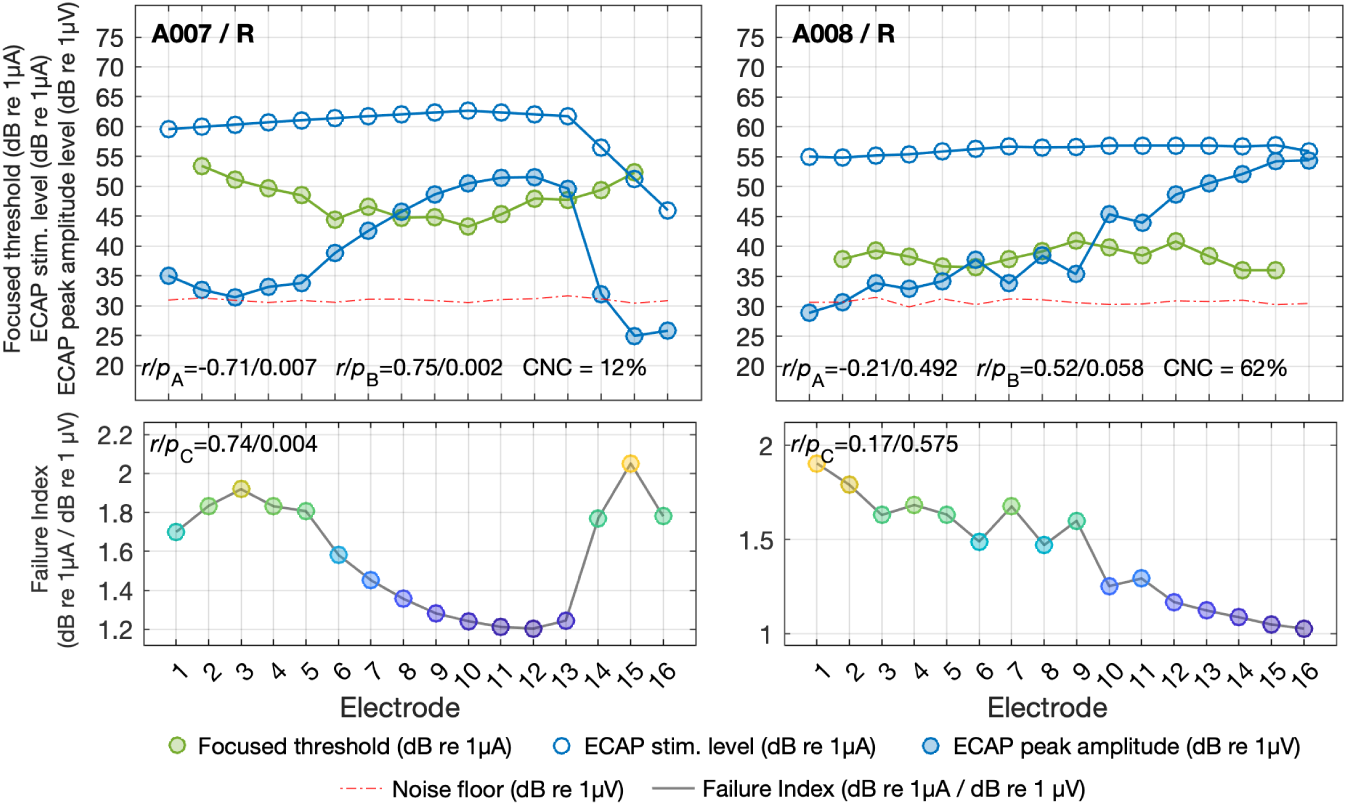
Focused thresholds and ECAP peak amplitudes (top panels) and FIs (bottom panels) for two example participants, A007 / R (left panels) and A008 / R (right panels); Electrodes on the x-axis, Focused thresholds (dB re 1 µA, green filled circles), ECAP stimulation current (dB re 1 µA, blue open circles), ECAP peak amplitudes (dB re 1 µV, blue filled circles), and ECAP noise floor (3*σ* of ECAP forward-masking noise frame, dB re 1 µV, dashed red line) on the y-axis; Partial correlation outcomes between focused thresholds and ECAP peak amplitudes (*r/p*_A_), between ECAP stimulation levels and peak amplitudes (*r/p*_B_), and CNC scores, are shown per individual. Failure Index across all available CI electrodes (bottom panels) color coded from low (blue) to high (yellow). Correlation outcomes between focused thresholds and FIs are shown (*r/p*_C_). Focused thresholds, ECAP data, and FIs of all 29 participants are presented in the Supplementary Fig. 1 and 2, and correlation outcomes in Supplementary Table 1.

Fig. 2 (bottom panels) shows the FI calculated from the ratio between ECAP input stimulation current levels and output peak amplitudes (color coded between the individual FI range for a clear representation, low FIs in blue to high FIs in yellow). A007 / R shows a positive relationship between focused thresholds and FI (*r* _C_ = .74, *p*_C_ = .004), with electrodes showing low focused detection threshold and low corresponding FIs — input-to-output ratio with ECAP amplitudes following stimulation current — and vice versa. Both metrics, therefore, seem to similarly detect changes of the ENI and the neural sensitivity to electric stimulation. On the contrary, A008 / R again does not show a clear relationship between the outcome measures.

Focused threshold measurements and ECAP peak amplitudes are highly variable and differ in shape and average level within and across all study participants (Supplementary Fig. 1). These variabilities are also reflected by the calculations of the FI (Supplementary Fig. 2). ECAP stimulation current levels show overall less variability across electrodes, but comparably large average differences across study participants.

### 3.1 Focused perceptual threshold and ECAP peak amplitude

In an initial analysis, we compared individual focused perceptual thresholds to ECAP peak amplitudes across the electrode array (Fig. 3, panel a and Supplementary Table 1). Linear correlations were performed and Pearson correlation coefficients calculated. ECAP peak amplitude data below noise (3*σ* of ECAP forward-masking noise frame) was removed before the analysis (n = 21, 5.17%) in addition to missing measurements (n = 11, 2.71%). The analysis showed large individual variations, with overall 15 individuals showing negative trends, 6 of them significant (A007 / R, A015 / L, A019 / R, A022 / L, A028 / R, A040 / L). A total of 14 individuals showed positive trends, but only one of them significant (A027 / L). Pearson correlation analysis across the averaged individual outcomes (*r* _mean_ = .250, *p*_mean_ = .187) shows no clear trend. An investigation of the electrode location (predictor variable) from apex-to-base as an influencing factor on the relationship between focused thresholds and ECAP peak amplitudes (response variables) via linear regression revealed, that electrode location explained a substantial portion of variance (*R^2^* values with *p* ă .05) for 18 individuals for at least one of the response variables, which is more than half of the study population. To account for this influence of electrode location on the correlation between focused thresholds and ECAP peak amplitudes controlling for electrode location, a follow-up analysis with partial correlations was performed. Overall, 7 individuals we found either significant partial correlations after controlling for electrode location, indicating that electrode order suppresses the true association, or a shift to non-significant partial correlations outcomes, indicating that the association was largely attributable to their mutual relationship with electrode location, mostly seen for the same individuals with large *R^2^* values before. Nevertheless, electrode location had no overall influence for the majority of individuals and for the investigated measures, expressed by either no or significant outcomes in both Pearson and the partial correlations.

**Figure 3:**
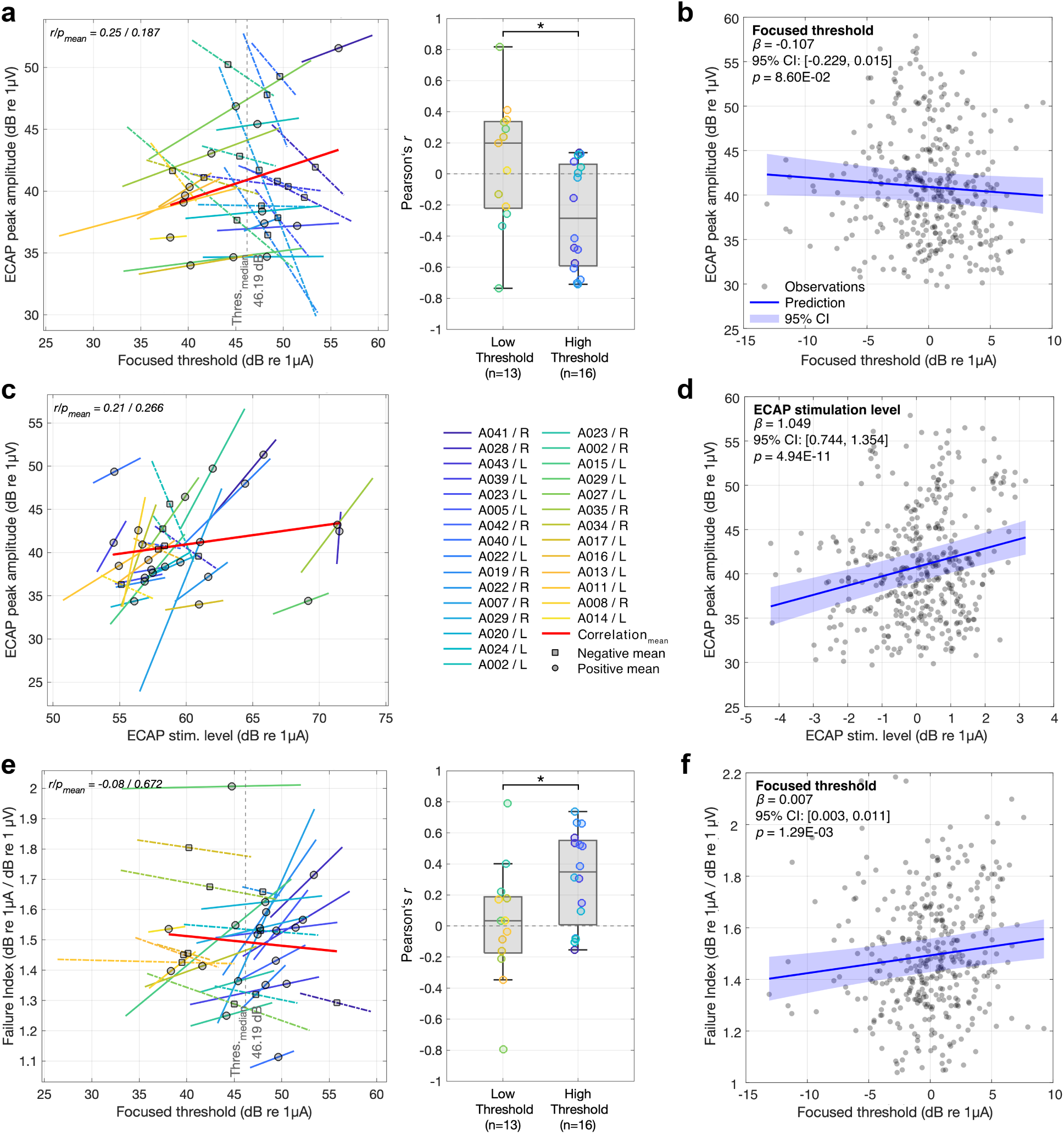
Panels a, c, and e show Pearson correlations analyses between focused threshold and ECAP peak amplitude (panel a, electrodes 2 to 15), ECAP stimulation current and peak amplitude (panel c, electrodes 1 to 16), and focused threshold and FI (panel e, electrodes 2 to 15). Correlation parameters on the x-axes and y-axes. Positive and negative correlations are indicated via solid and dotted lines (color codes from blue to yellow represent participants sorted from high to low average focused thresholds), and circle and square average values across electrodes for both correlation variables (in gray), respectively. Pearson correlation analyses across all study participants and average outcomes (indicated in red) were not significant. The box plots of panel a and panel e show Pearson correlation coefficients median-split at focused thresholds of 46.19 dB re 1 µA, indicated as vertical dashed lines. The full list of correlation outcomes between the investigated variables is listed in Supplementary Table 1. Panels b, d, and f show LMM outcomes for the investigated models (Eq. 3, 4, 5); Panel b shows the LMM predictions (Eq. 3) for focused perceptual thresholds (dB re 1 µA, x-axis) and ECAP peak amplitudes (dB re 1 µV, y-axis). Panel d shows the LMM predictions (Eq. 4) for ECAP stimulation current levels (dB re 1 µA, x-axis) and ECAP peak amplitudes (dB re 1 µV, y-axis). Panel f shows the LMM predictions (Eq. 5) for focused thresholds (dB re 1 µA, x-axis) and FI (dB re 1 µA { (dB re 1 µV, y-axis).

The box plot of Fig. 3, panel a shows Pearson correlation coefficients between focused perceptual threshold and ECAP peak amplitude grouped by low and high thresholds. Groups were defined by a median-split of focused thresholds at 46.19 dB re 1 µA. A shift from slightly positive Pearson correlation outcomes with a low-threshold group median of *r* = .198, to a negative high-threshold group median of *r* = –.285 can be seen. Despite substantial variabilities in Pearson correlation coefficients, the differences between the median-split groups were statistically significant (*Z* = 2.26, *p* = .024).

To improve generalizability of these findings beyond the individual relationships and the consideration of electrode location, a LMM was fitted with ECAP peak amplitude as the dependent variable, focused threshold as fixed effect, and subject as random intercept to control for repeated measures and subject variability (Eq. 3). Model predictions are shown in Fig. 3, panel b. Focused thresholds and the ECAP peak amplitudes show a small but not significant negative association (*β* = –0.107, *t* (372) = –1.725, *p* = .086). Hence, focused thresholds might be sensitive to objective ECAP measurements across electrodes represented by the peak amplitudes (higher focused thresholds at lower ECAP peak amplitudes and vice versa), however, we did not see a strong relationship. Also, the variance explained by the fixed effect focused threshold was neglectable (marginal *R^2^* = .004), and the full model including random intercepts captures most of the variance (53.3%) between focused thresholds and ECAP peak amplitudes (conditional *R^2^* = .533).

### 3.2 ECAP stimulation current and ECAP peak amplitude

We extended our investigation of the ENI and fitted a LMM with ECAP peak amplitudes as dependent variable, ECAP stimulation current as a fixed effect, and subject as random intercept (Eq. 4). Similar to the first model, ECAP peak amplitude data below noise (n = 30, 6.47%) and missing data was removed (n = 22, 4.74%). This model shows the association of ECAP input and output. Similar to the initial analysis we performed individual correlations beforehand (Fig. 3, panel c), resulting in a positive correlation for the majority of 23 individuals, 9 of who were significant (A002 / R, A007 / R, A015 / L, A022 / L, A022 / R, A027 / L, A034 / R, A035 / R, A041 / R). Six individuals showed an opposite linear relationship, with only two of them significant (A002 / L, A023 / R). Correlation outcomes are shown in Supplementary Fig. 1 and Supplementary Table 1. The Pearson correlation analysis across the averaged outcomes (*r* _mean_ = .21, *p*_mean_ = .266) was not significant with no clear trend, influenced by three participants (A028 / R, A029 / L, A035 / R) with much higher stimulation levels than the rest of the study population.

The model demonstrated a strong, statistically significant positive association between ECAP stimulation current level and peak amplitudes across all study individuals (*β* = 1.049, *t* (410) = 6.754, *p* ă .0001), seen in Fig. 3, panel d, indicating that increases in ECAP stimulation levels are associated with substantial increases of ECAP peak amplitudes. Hence, stimulation level might be a possible driving force in the efficacy of electric stimulation at the ENI and the resulting neural response. Nevertheless, the fixed effect ECAP stimulation level alone explained only 5.0% of the variance in the outcome (marginal *R^2^* = .050), while the full model including random intercepts explained 57.9% of the variance (conditional *R^2^* = .579). Hence, also here a large proportion of variance was attributable to differences between individuals.

### 3.3 Focused perceptual thresholds and the Failure Index

An evaluation of focused perceptual thresholds and the FI across subjects via Pearson correlations (Fig. 3, panel e) showed, that the majority of 19 individuals followed a positive correlation, 5 of them were significant (A007 / R, A015 / L, A019 / R, A022 / L, A028 / R), and similar to the previous comparison of focused thresholds and ECAP peak amplitudes, A027 / L showed a significant opposite relationship. The Pearson correlation analysis across the averaged individual outcomes (*r* _mean_ = –.08, *p*_mean_ = .672) was not significant with no clear trend, similar as we saw it for focused thresholds and ECAP peak amplitudes.

The median-split of focused thresholds showed a shift in correlation coefficients for focused perceptual threshold and FI (box plot of Fig. 3, panel e). While the low-threshold group shows Pearson correlation coefficients close to zero (*r* = .034), the high-threshold group results in mostly positive trends (*r* = .349). Also, these group differences were statistically significant (*Z* = –2.04, *p* = .041).

To investigate the effects across our study population, we defined a third LMM (Eq. 5) comprising focused thresholds (fixed effect) and the FI (dependent variable), with subjects as random intercept. Data below noise (n = 21, 5.17%) and missing data (n = 11, 2.71%) was removed. We found a statistically significant positive associations between focused thresholds and the FIs (*β* = .007, *t* (372) = 3.241, *p* = .001), shown in Fig. 3, panel f. Even though only approximately 1.2% of the FI’s variance was explained via the focused threshold (marginal *R^2^* = .012), and subject variability adds most of the variance with approximately 59.6% again (conditional *R^2^* = .596), these findings indicate that increasing focused thresholds are associated with increasing FIs, a finding which suggests that high focused thresholds might indicate areas of decreased ENI quality or neural sensitivity, represented by lower ECAP amplitude and higher ECAP stimulation currents.

### 3.4 Effect of Speech perception and CI experience

As a last step, we tested whether the effects of clinical speech performance and CI listening experience on the FI vary depending on the level of focused perceptual thresholds. Therefore, we added speech and CI experience to the previous LMM (Eq. 6) and performed the analysis considering missing CNC score of one study participant (3.45% of the data). Co-linearity of CNC scores and CI experience was ruled out, as Kendall’s *τ* revealed no significant correlation between the two variables (*τ* = .223, *p* = .104). Similar to the simpler model (Eq. 5), the extended LMM showed a strong positive main effect for focused thresholds (*β* = .036, *t* (354) = 4.178, *p* ă .0001), indicating that higher threshold values were associated with higher FIs (Fig. 4). Importantly, the interaction between focused thresholds and speech perception showed a significant effect in predicting the FI (Focused thresholds ^ CNC score: *β* = –4.82E-04, *t* (354) = –3.766, *p* = .0002). In other words, the positive association between focused thresholds and FIs was stronger in individuals with lower speech scores. For a better representation, CNC scores were split at the population median (CNC_median score_ = 70%, color coded for low and high speech performance), a similar method to our previous work (e.g., [45, 46]). Furthermore, neither the main effects of speech (*β* = 2.92E-04, *t* (354) = 0.158, *p* = .875) or CI experience (*β* = –.0015, *t* (354) = –.236, *p* = .814), nor their interaction with thresholds (Focused thresholds ^ CI experience: *β* = 7.61E-04, *t* (354) = 1.470, *p* = .143) reached statistical significance. In comparison to the previous model, including CNC performance and CI experience accounted for more variance through fixed effects (marginal *R^2^* = .031) and also showed a slight increase in overall explained variance (conditional *R^2^* = .622), indicating a somewhat better fit. However, the overall variance explained by the fixed effects is still low, meaning the predictors of this model only modestly explain their association to the FIs, and there are unknown factors not addressed in the current analysis.

**Figure 4:**
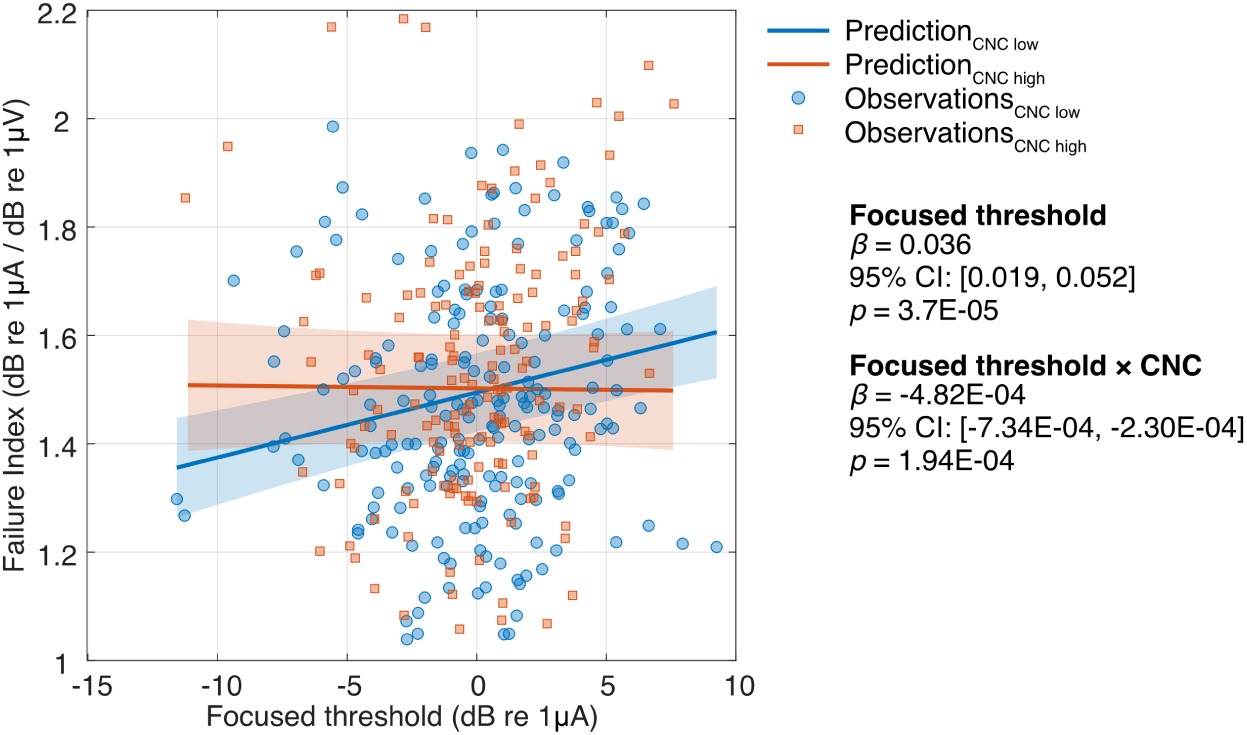
Represents the LMM predictions for the expanded FI model (Eq. 6) and the interaction of focused thresholds and clinically assessed CNC score. This should highlight the relationship between focused thresholds and FIs of two speech performance groups, median-split at 70% into low and high CNC performance. The low performance group (CNC scores ď 70%) shows a strong positive relationship between focused threshold and FI, while the high performance group (CNC scores ą 70%) does not show an effect.

All above presented LMM outcomes are listed in Table 2. Low marginal and much higher conditional *R^2^* values indicted a minimal contribution from the different fixed effects, while random effects accounted for nearly all of the variance. Hence, only an overall small amount of the variance could be explained by investigating the relationships between focused thresholds, ECAP stimulation current level and peak amplitudes, as well as clinical CNC scores, and CI experience in terms of related measures for neural sensitivity.

**Table 2:** Linear mixed-effects models and parameter estimates (*β*), standard errors (SE), test statistic (*t*), *p*-value, 95% lower and upper confidence intervals (CI) around the estimate, and the model marginal and conditional *R^2^*.

| Model | Predictor | Estimate ($\beta$) | Standard Error (SE) | $t$ | $p$ | 95% CI (lower, upper) |
| --- | --- | --- | --- | --- | --- | --- |
| **ECAP peak amplitude ~ Focused threshold + (1\|subject)** |  |  |  |  |  |  |
|  | (Intercept) | 40.91 | 0.8543 | 47.889 | < **0.001** | 39.23, 42.589 |
|  | Foc. threshold | -0.1067 | 0.0620 | -1.721 | 0.0860 | -0.229, 0.015 |
| $R^2$ marginal / conditional: [0.004, 0.533] | | | | | | |
| **ECAP peak amplitude ~ ECAP stimulation level + (1\|subject)** |  |  |  |  |  |  |
|  | (Intercept) | 40.772 | 0.8583 | 47.503 | < **0.001** | 39.085, 42.460 |
|  | ECAP stim. level | 1.0491 | 0.1553 | 6.754 | < **0.001** | 0.744, 1.354 |
| $R^2$ marginal / conditional: [0.050, 0.579] | | | | | | |
| **Failure Index ~ Focused threshold + (1\|subject)** |  |  |  |  |  |  |
|  | (Intercept) | 1.493 | 0.0329 | 45.379 | < **0.001** | 1.428, 1.558 |
|  | Foc. threshold | 0.0069 | 0.0021 | 3.242 | **0.0013** | 0.0027, 0.0111 |
| $R^2$ marginal / conditional: [0.012, 0.596] | | | | | | |
| **Failure Index ~ Focused threshold × CNC score + Focused threshold × CI experience + (1\|subject)** |  |  |  |  |  |  |
|  | (Intercept) | 1.4867 | 0.1170 | 12.712 | < **0.001** | 1.257, 1.717 |
|  | Foc. threshold | 0.0355 | 0.0085 | 4.179 | < **0.001** | 0.0188, 0.0522 |
|  | CNC score | 2.92E-04 | 0.0019 | 0.158 | 0.875 | -0.00335, 0.00393 |
|  | CI experience | -0.0015 | 0.0064 | -0.236 | 0.813 | -0.0141, 0.0111 |
|  | Foc. threshold : CNC score | -4.82E-04 | 1.28E-04 | -3.766 | < **0.001** | -7.34E-04, -2.30E-04 |
|  | Foc. threshold : CI experience | 7.61E-04 | 5.18E-04 | 1.470 | 0.143 | -2.58E-04, 0.00178 |
| $R^2$ marginal / conditional: [0.031, 0.622] | | | | | | |

## 4 Discussion

Neural sensitivity to electric stimulation in cochlear implants derived from physiological ECAP recordings and focused perceptual thresholds to estimate their relationship were explored. In addition to individual correlations, four LMMs comprising different combinations of model parameters were investigated.

The individual investigation of ECAP peak responses and focused perceptual thresholds where mixed throughout our study population, similar to previously reported findings [4, 18, 39]. DeVries et al. [4] showed a similarly weak but significant negative relationship on the group level. Garcia et al. [18] compared normalized focused threshold measurements to their neural health estimate derived from panoramic-ECAP recordings, and saw a similar relationship by correlating the two perceptual and electrophysiological metrics. Peng et al. [39] compared perceptual threshold variability derived from monopolar and bipolar threshold measurements with ECAP peak response amplitudes, and also saw a weak but significant relationship between the two measures. While we did see larger variabilities across subjects, we could confirm previous findings, when we accounted for subject variability. Our first LMM (Eq. 3) showed an inverse relationship between focused thresholds and ECAP peak amplitudes on the group level. However, large variabilities in both outcome measures led to very low explanatory power of the investigated model with most variance covered by individual variability. A possible explanation of this finding is that the dependency of focused thresholds on electrode placement in the cochlea and the distance to the modiolus might both increase focused thresholds and reduce response amplitudes [34, 37]. Overall, different perceptual threshold measurements, either directly assessed via the threshold itself or derived from the difference between monopolar and bipolar thresholds, and physiological measurements seem to capture the same aspects of the ENI, indicating some weak relationships to the sensitivity of the auditory neurons.

In an individual comparison of ECAP stimulation current and ECAP peak amplitudes (Fig. 3, panel c) we observed a strong dependency between the individual input and output for most individuals of our study population. However, ECAP peak amplitudes along the CI electrode array do not always follow the ECAP stimulation current. While the stimulation current does not vary much between neighboring electrodes, possibly attributable to the monopolar stimulation mode and wide current spread reaching a large population of auditory neurons, the objective measurable ECAP response to a specific stimulus and electrode location shows large variability, possibly indicating differences in local neural activity. Simple correlations between ECAP input and output might therefore be insufficient, to capture the actual variation of neural sensitivity to electric stimulation along the electrode array. Similar to these individual comparisons, our LMM (Eq. 4) was not able to capture the subtle differences across people and electrodes, resulting in a significant positive relationship on the group level.

We observed an interesting relationship of focused thresholds and both ECAP peak amplitudes and FIs (box plots of Fig. 3, panels a and e). Subjects with focused behavioral thresholds higher than 46.19 dB (the median for these 29 ears) show a strong correspondence between focused threshold and ECAP peak amplitudes and FI, whereas those with lower focused thresholds did not. Because we also observe that subjects with lower focused thresholds tend to have better speech perception, this relationship could suggest that the variability in focused thresholds for the better performers (with lower overall thresholds) may be a result of relatively poor ENIs from some other factor than neural degeneration. Other factors could include greater distance between electrodes and target neurons [34, 37] or bone and tissue growth that may impede the flow of current to neurons, among others.

To better address the variabilities between ECAP stimulation current and ECAP peak amplitudes, a new metric that may be sensitive to neural degeneration, called the failure index (FI), was first validated in an animal model [19]. We could confirm these initial findings from the animal model for FIs derived from the ECAP parameters in our subject population, by calculating the FI per electrode and within subject (Eq. 2, examples in Fig. 2, bottom panels, and Supplementary Fig. 2). To investigate, if neural sensitivity to electric stimulation in humans can be captured more robustly by combining focused thresholds and ECAP stimulation levels and peak amplitudes through the FI, we combined the two measures into a LLM (Eq. 5). The model estimated a positive relationship between the outcome metrics, however, similar to the LMM comparing ECAP peak amplitudes and focused thresholds (Eq. 3), the explanatory power was low with only 1.2% of the variance explained by focused thresholds, and most variance explained by large variability among CI individuals. Garcia et al. [29] mentioned the electrode-to-modiolus distance estimated from CT scans as a possible influential factor, showing that FI estimates might be biased by the electrode placement in the cochlea. In contrast to Garcia et al. [29], we recorded the ECAP at higher current levels rather than at MCL. Our recording level at a rating of 7 “loud but comfortable” might get closer to the ECAP saturation level as was used in the guinea pig measures by Konerding et al. [19]. Hence, this might counteract the impacts of electrode-to-modiolus distance on FI, further supported by findings of the Konerding study, that the FI is robust down to about 70% of the input dynamic range. However, more evidence in terms of a broader evaluation of the ENI and electrode placement in human CI subjects is needed to further support this statement.

The introduction of CI experience and clinically assessed CNC scores to the model (Eq. 6) revealed, that the relationship between focused thresholds and FI differs for CI individuals depending on their level of clinical speech performance. We used a median split to divide participants at 70% performance and found that individuals with poor clinical speech performance have a stronger relationship between the two outcome measures, while people with good speech performance show weaker effects, and that there was no influence of CI experience. Previously, several studies showed a somewhat comparable finding, a negative correlation between focused threshold and clinical CNC performance scores, with high focused thresholds corresponding to low CNC performance, and vice versa [4, 34, 37]. Similarly, Peng et al. [39] confirmed this for the variabilities in the difference between monopolar and bipolar thresholds. These outcomes might indicate the importance of good neural sensitivity for good speech perception outcomes, with low thresholds and ECAP peak amplitudes following the ECAP stimulation current, i.e. low FIs, across the whole CI array. For people with high speech performance scores, on the contrary, the weaker relationship between ECAP stimulation current and peak amplitudes or focused perceptual thresholds as neural estimates seems to be of less importance. In other words, CI individuals with better speech performance might be less affected by variabilities in their neural activity in CI stimulation across electrodes, and poor ENIs might originate in other factors such as electrode-to-modiolus distance. Taken together, these findings suggest that the quality of the ENI or neural sensitivity derived from human ECAP recordings and focused threshold measures drive the overall speech performance capabilities of CI listeners.

Novel bio-inspired CI coding strategies have investigated possibilities to compensate for the variability of the ENI. For example, focused stimulation was proposed as promising alternative to individualize CI stimulation tailored to the variability of the ENI (e.g., [35, 37, 47–50]). In addition, deactivating channels based on suboptimal signal transmission defined by perceptual drawbacks in electrode discrimination, temporal modulation sensitivity or physiological IPG outcomes, has shown some promising results (e.g., [51–54]. Bierer and Litvak [45] implemented a coding strategy combining channel deactivation based on high focused thresholds and partial tripolar (pTP) stimulation, an electrode configuration comprised of an electrode triplet with its flanking electrodes being phase inverted to return parts of the stimulation current and a fraction of the current via an extra cochlear ground electrode. CI stimulation with the pTP configuration led to improvements in speech perception performance, especially for clinically poor performers. Similar findings were presented by Arenberg et al. [55], who showed that a novel strategy using dynamic tripolar (DT) stimulation, where a proportion of the stimulating current being dynamically adjusted based on stimulation level in addition to the pTP mode, led to an improvement in speech perception. Wohlbauer et al. [46] combined channel deactivation based on high focused thresholds and DT stimulation into one optimized strategy and showed benefits for the novel CI coding strategy for some individual participants. The current findings further highlight the importance of CI listeners’ neural sensitivity on new CI stimulation strategies, and performance improvements could be expected if the ENI and neural sensitivity to electric stimulation is understood in more depth.

## 5 Conclusion

In the current study, perceptual and physiological neural health measures were investigated on an individual level via linear correlations and in a number of LLMs to assess the relation to neural sensitivity. We showed that focused thresholds, ECAP stimulation current, and ECAP peak amplitudes are indeed linked to each other. Overall, our findings suggest, that the sensitivity of the auditory neurons can be estimated by focused thresholds and objective ECAP recordings. Furthermore, the relationship between ECAP stimulation level and ECAP peak amplitudes, i.e. the FI, revealed the large variability in the ECAP input-to-output relationship in human CI listeners. These findings further highlight the potential of the individual evaluation of the ENI with a combination of focused thresholds and ECAP parameters as an early estimate the speech performance abilities of that subject.

Nevertheless, the explored models could only explain a small proportion of the variance of the findings, which highlights, that additional factors such as electrode position in the cochlea may play a crucial role in these relationships. Including imaging data, for example, to subtract the interacting component of CI array insertion and placement might be a necessary step to increase the explanatory power and should be considered in future analyses.

## Data Availability

All data produced in the present study are available upon reasonable request to the authors.

## Acknowledgements

We sincerely thank all cochlear implant participants for their time and commitment to this study. We thank Scott Aker, Faten Awwad, Charles B. Hem, Caylin McCallick for their support during data collection and participant recruitment.

## Supplementary information

The online version contains supplementary material available at the end of the document.

## Ethics Approval and Consent to Participate

The current study was approved by the institutional review boards (IRBs) at Massachusetts Eye and Ear. All study participants provided written informed consent before participation in the study.

---

**Supplementary Figure 1:**
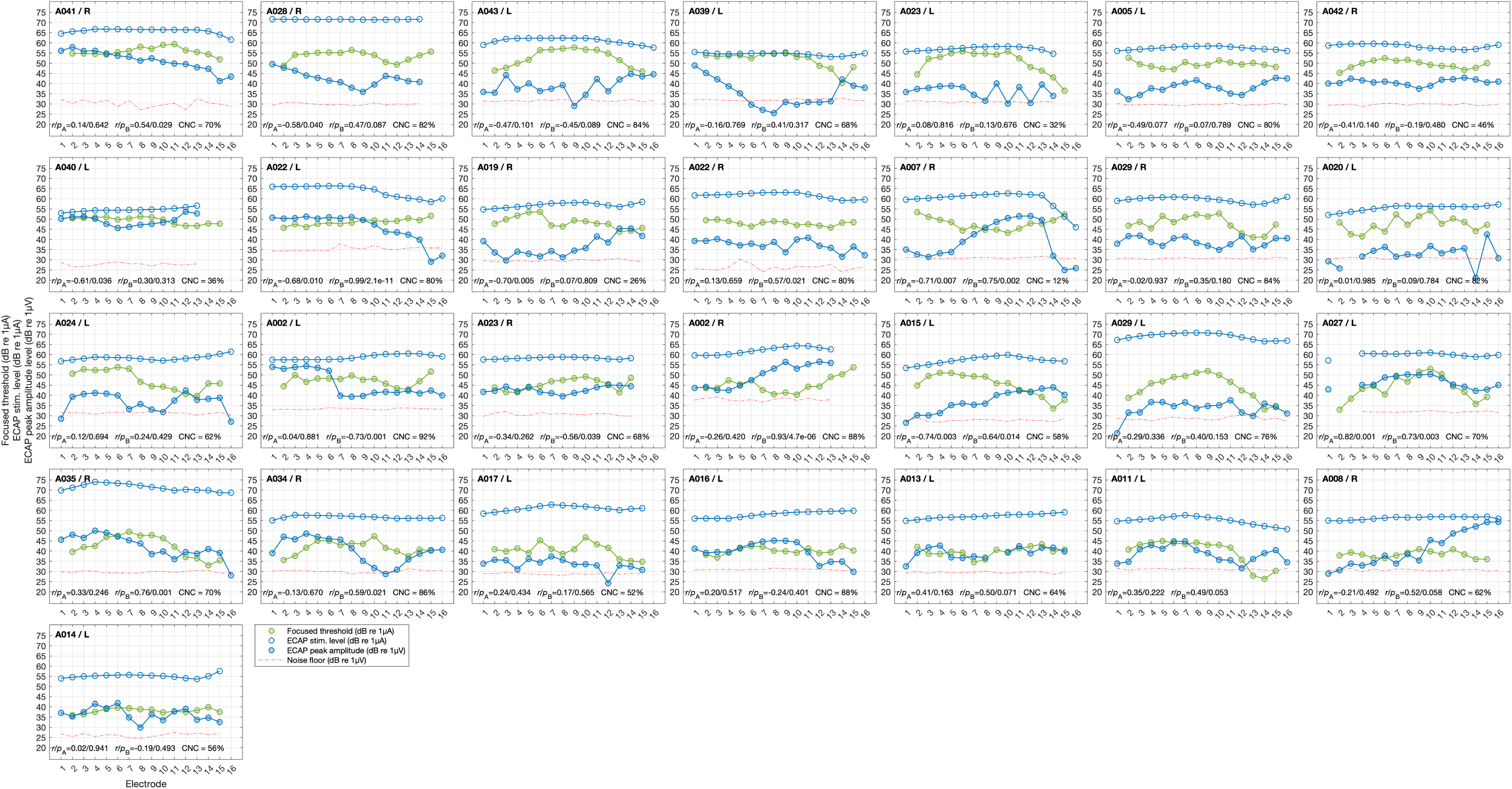
Focused thresholds (green filled circles), electrically evoked compound action potential (ECAP) stimulation current (blue open circles) and peak amplitudes (blue filled circles) from all study participants across all available CI electrodes. Pearson correlation results between focused thresholds and ECAP peak amplitudes, and between ECAP simulation current levels and ECAP peak amplitudes, and consonant-nucleus-consonant (CNC) scores, are shown per individual.

**Supplementary Figure 2:**
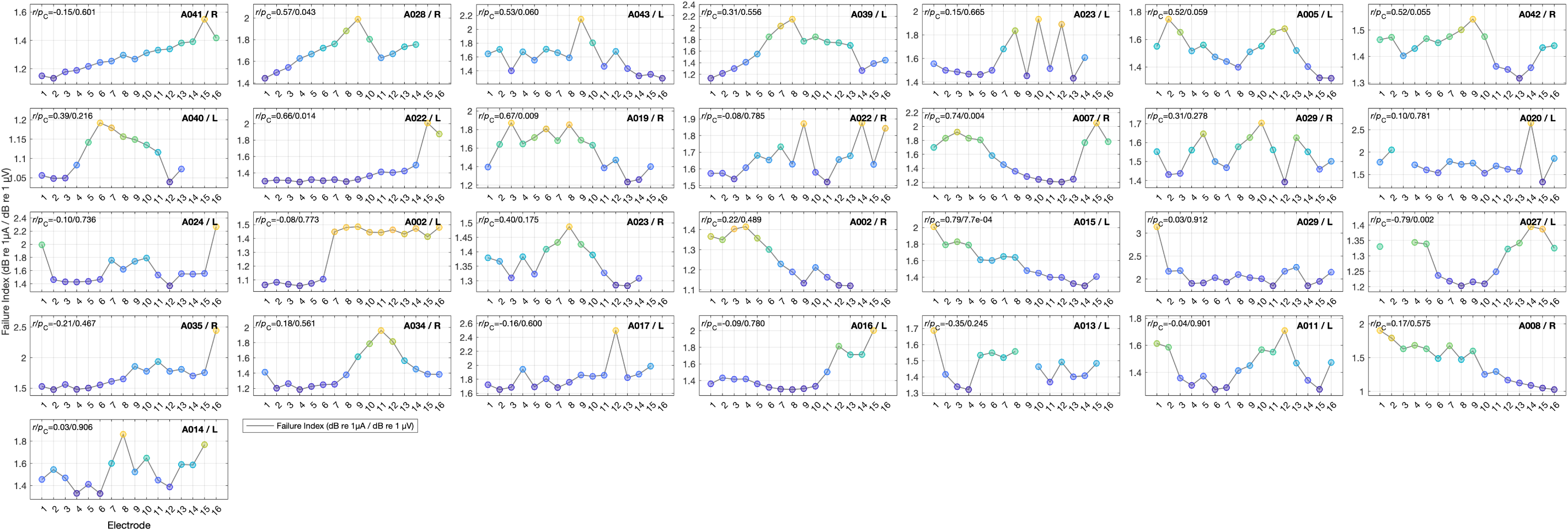
Failure Index (FI) for all study participants across all available cochlear implant electrodes, and the Pearson correlation between focused threshold and Failure Index (FI). FI color coded from low (blue) to high (yellow). Participants are sorted from high to low focused thresholds, averaged across all electrodes, to align with our previous work.

**Supplementary Table 1:**
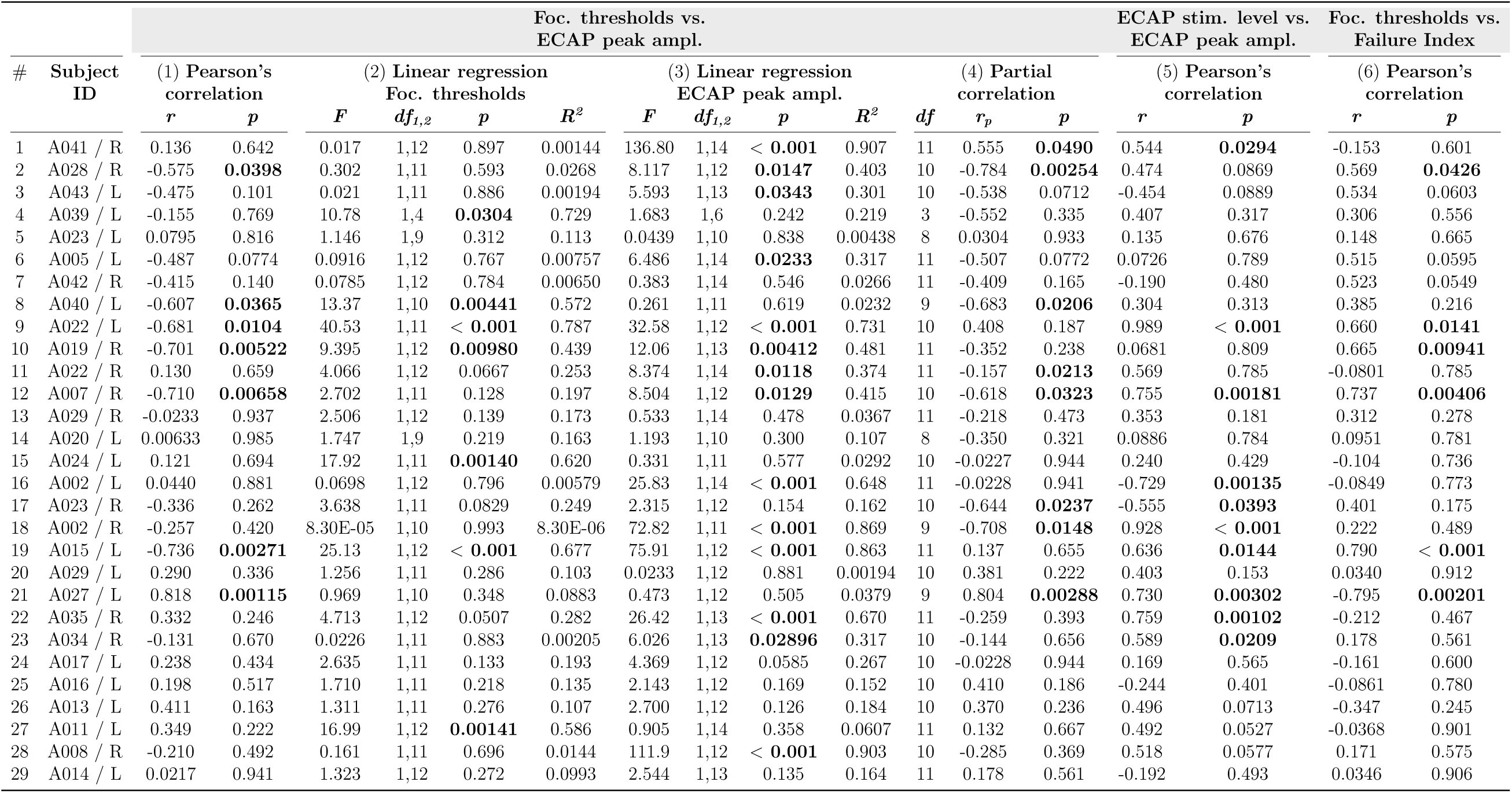
Summary of initial analysis for all subjects; (1) Pearson’s correlations between focused thresholds and ECAP peak amplitudes, (2) linear regression between focused thresholds and electrodes, (3) linear regression between ECAP peak amplitudes and electrodes, (4) Partial correlations between focused thresholds and ECAP peak amplitudes, (5) Pearson’s correlations between ECAP stimulation levels and ECAP peak amplitudes, (6) Pearson’s correlations between focused thresholds and the Failure Index.

## References

[1] Zeng FG. 2022. Celebrating the one millionth cochlear implant. JASA Express Letters 2(7):077201

[2] Aschendorff A, Kromeier J, Klenzner T, Laszig R. 2007. Quality control after insertion of the nucleus contour and contour advance electrode in adults. Ear and Hearing 28(SUPPL.2):75–79

[3] Finley CC, Holden TA, Holden LK, Whiting BR, Chole RA, et al. 2008. Role of electrode placement as a contributor to variability in cochlear implant outcomes. Otology and Neurotology 29(7):920–928

[4] DeVries L, Scheperle R, Bierer JA. 2016. Assessing the Electrode-Neuron Interface with the Electrically Evoked Compound Action Potential, Electrode Position, and Behavioral Thresholds. JARO – Journal of the Association for Research in Otolaryngology 17(3):237–252

[5] Wilson B, Finley C, Weber B, White M, JC FJ, et al. 1988. Comparative studies of speech processing strategies for cochlear implants. The Laryngoscope 98(10):1069–1077

[6] Skinner MW, Holden LK, Whitford LA, Plant KL, Psarros C, Holden TA. 2002. Speech recognition with the Nucleus 24 SPEAK, ACE, and CIS speech coding strategies in newly implanted adults. Ear and Hearing 23(3):207–223

[7] Wohlbauer DM, Dillier N. 2025. A Hundred Ways to Encode Sound Signals for Cochlear Implants. Annual Review of Biomedical Engineering 27(1):335–369

[8] Otte J, Schuknecht H, Laryngoscope AKT, 1978 U. 1978. Ganglion cell populations in normal and pathological human cochleae. Implications for cochlear implantation. Wiley Online Library 88(8):1252–1267

[9] Nadol JB, Young yS, Glynn RJ. 1989. Survival of spiral ganglion cells in profound sensorineural hearing loss: implications for cochlear implantation. *Annals of Otology*, Rhinology, and Laryngology 98(6):411–416

[10] Makary CA, Shin J, Kujawa SG, Liberman MC, Merchant SN. 2011. Age-related primary cochlear neuronal degeneration in human temporal bones. JARO – Journal of the Association for Research in Otolaryngology 12(6):711–717

[11] Kujawa SG, Liberman MC. 2006. Acceleration of age-related hearing loss by early noise exposure: Evidence of a misspent youth. Journal of Neuroscience 26(7):2115–2123

[12] Kujawa SG, Liberman MC. 2009. Adding insult to injury: Cochlear nerve degeneration after ”temporary” noise-induced hearing loss. Journal of Neuroscience 29(45):14077–14085

[13] Smith L, Simmons FB. 1983. Estimating eighth nerve survival by electrical stimulation. *Annals of Otology*, Rhinology and Laryngology 92(1):19–23

[14] Shepherd RK, Javel E. 1997. Electrical stimulation of the auditory nerve. I. Correlation of physiological responses with cochlear status. Hearing Research 108(1-2):112–144

[15] Prado-Guitierrez P, Fewster LM, Heasman JM, McKay CM, Shepherd RK. 2006. Effect of interphase gap and pulse duration on electrically evoked potentials is correlated with auditory nerve survival. Hearing Research 215(1-2):47–55

[16] Ramekers D, Versnel H, Strahl SB, Smeets EM, Klis SF, Grolman W. 2014. Auditory-nerve responses to varied inter-phase gap and phase duration of the electric pulse stimulus as predictors for neuronal degeneration. JARO – Journal of the Association for Research in Otolaryngology 15(2):187–202

[17] Cosentino S, Gaudrain E, Deeks JM, Carlyon RP. 2015. Multistage nonlinear optimization to recover neural activation patterns from evoked compound action potentials of cochlear implant users. IEEE Transactions on Biomedical Engineering 63(4):833–840

[18] Garcia C, Goehring T, Cosentino S, Turner RE, Deeks JM, et al. 2021. The Panoramic ECAP Method: Estimating Patient-Specific Patterns of Current Spread and Neural Health in Cochlear Implant Users. JARO – Journal of the Association for Research in Otolaryngology 22(5):567–589

[19] Konerding W, Arenberg J, Sznabel D, Kral A, Baumhoff P. 2025. An Electrically Evoked Compound Action Potential Marker for Local Spiral Ganglion Neuron Degeneration: The Failure Index. Journal of Neuroscience 45(7)

[20] Abbas PJ, Brown CJ, Shallop JK, Firszt JB, Hughes ML, et al. 1999. Summary of results using the nucleus CI24M implant to record the electrically evoked compound action potential. Ear and Hearing 20(1):45–59

[21] Wai Kong Lai, Dillier N. 2000. A simple two-component model of the electrically evoked compound action potential in the human cochlea. Audiology and Neuro-Otology 5(6):333–345

[22] Cohen LT, Richardson LM, Saunders E, Cowan RS. 2003. Spatial spread of neural excitation in cochlear implant recipients: Comparison of improved ECAP method and psychophysical forward masking. Hearing Research 179(1-2):72–87

[23] Abbas PJ, Hughes ML, Brown CJ, Miller CA, South H. 2004. Channel interaction in cochlear implant users evaluated using the electrically evoked compound action potential. Audiology and Neuro-Otology 9(4):203–213

[24] Schvartz-Leyzac KC, Colesa DJ, Swiderski DL, Raphael Y, Pfingst BE. 2023. Cochlear Health and Cochlear-implant Function. JARO – Journal of the Association for Research in Otolaryngology 24(1):5–29

[25] McKay CM, Henshall KR. 2003. The perceptual effects of interphase gap duration in cochlear implant stimulation. Hearing Research 181(1-2):94–99

[26] He S, Xu L, Skidmore J, Chao X, Riggs WJ, et al. 2020. Effect of Increasing Pulse Phase Duration on Neural Responsiveness of the Electrically Stimulated Cochlear Nerve. Ear and Hearing 41(6):1606–1618

[27] Brochier T, Guérit F, Deeks JM, Garcia C, Bance M, Carlyon RP. 2021. Evaluating and Comparing Behavioural and Electrophysiological Estimates of Neural Health in Cochlear Implant Users. JARO – Journal of the Association for Research in Otolaryngology 22(1):67–80

[28] Hughes ML. 2008. A re-evaluation of the relation between physiological channel interaction and electrode pitch ranking in cochlear implants. The Journal of the Acoustical Society of America 124(5):2711–2714

[29] Garcia C, Sismono F, Goehring T, Guerit F, Arzounian D, Carlyon RP. 2025. A comparison of electrophysiological measures for characterizing the cochlear-implant electrode-neuron interface. JASA Express Letters 5(8):1–8

[30] Garcia C, Carlyon RP. 2025. Assessing Array-Type Differences in Cochlear Implant Users Using the Panoramic ECAP Method. Ear and Hearing 46(5):1355–1368

[31] Zhou N, Pfingst BE. 2014. Relationship between multipulse integration and speech recognition with cochlear implants. The Journal of the Acoustical Society of America 136(3):1257–1268

[32] Zhou N, Dong L. 2017. Evaluating multipulse integration as a neural-health correlate in human cochlear-implant users: Relationship to psychometric functions for detection. Trends in Hearing 21:1–12

[33] Nelson DA, Donaldson GS, Kreft H. 2008. Forward-masked spatial tuning curves in cochlear implant users. The Journal of the Acoustical Society of America 123(3):1522–1543

[34] DeVries L, Arenberg JG. 2018. Psychophysical Tuning Curves as a Correlate of Electrode Position in Cochlear Implant Listeners. JARO – Journal of the Association for Research in Otolaryngology 19(5):571–587

[35] Bierer JA. 2007. Threshold and channel interaction in cochlear implant users: Evaluation of the tripolar electrode configuration. The Journal of the Acoustical Society of America 121(3):1642– 1653

[36] Bierer JA, Faulkner KF. 2010. Identifying cochlear implant channels with poor electrode-neuron interface: Partial tripolar, single-channel thresholds and psychophysical tuning curves. Ear and Hearing 31(2):247–258

[37] Long CJ, Holden TA, McClelland GH, Parkinson WS, Shelton C, et al. 2014. Examining the electro-neural interface of cochlear implant users using psychophysics, CT scans, and speech understanding. JARO – Journal of the Association for Research in Otolaryngology 15(2):293– 304

[38] Bierer JA, Bierer SM, Kreft HA, Oxenham AJ. 2015. A fast method for measuring psychophysical thresholds across the cochlear implant array. Trends in Hearing 19:1–12

[39] Peng T, Garcia C, Haneman M, Shader MJ, Carlyon RP, McKay CM. 2025. Comparing Patient-Specific Variations in Intra-Cochlear Neural Health Estimated Using Psychophysical Thresholds and Panoramic Electrically Evoked Compound Action Potentials (PECAPs). JARO – Journal of the Association for Research in Otolaryngology 26(1):77–91

[40] Scheperle RA, Abbas PJ. 2015. Relationships among peripheral and central electrophysiological measures of spatial and spectral selectivity and speech perception in cochlear implant users. Ear and Hearing 36(4):441–453

[41] Arslan NO, Luo X. 2022. Assessing the Relationship Between Pitch Perception and Neural Health in Cochlear Implant Users. JARO – Journal of the Association for Research in Otolaryngology 23(6):875–887

[42] Schvartz-Leyzac KC, Pfingst BE. 2018. Assessing the relationship between the electrically evoked compound action potential and speech recognition abilities in bilateral cochlear implant recipients. Ear and Hearing 39(2):344–358

[43] Langner F, Arenberg JG, Büchner A, Nogueira W. 2021. Assessing the relationship between neural health measures and speech performance with simultaneous electric stimulation in cochlear implant listeners. PLoS ONE 16(12 December)

[44] Nakagawa S, Schielzeth H. 2013. A general and simple method for obtaining R2 from generalized linear mixed-effects models. Methods in Ecology and Evolution 4(2):133–142

[45] Bierer JA, Litvak L. 2016. Reducing Channel Interaction Through Cochlear Implant Programming May Improve Speech Perception: Current Focusing and Channel Deactivation. Trends in Hearing 20:2331216516653389

[46] Wohlbauer DM, Hem CB, McCallick C, Arenberg JG. 2025. Speech performance in adults with cochlear implants using combined channel deactivation and dynamic current focusing. Hearing Research 463:109285

[47] Kral A, Hartmann R, Mortazavi D, Klinke R. 1998. Spatial resolution of cochlear implants: The electrical field and excitation of auditory afferents. Hearing Research 121(1-2):11–28

[48] Mens LH, Berenstein CK. 2005. Speech perception with mono– and quadrupolar electrode configurations: A crossover study. Otology and Neurotology 26(5):957–964

[49] van den Honert C, Kelsall DC. 2007. Focused intracochlear electric stimulation with phased array channels. The Journal of the Acoustical Society of America 121(6):3703

[50] Srinivasan AG, Padilla M, Shannon RV, Landsberger DM. 2013. Improving speech perception in noise with current focusing in cochlear implant users. Hearing Research 299:29–36

[51] Zwolan TA, Collins LM, Wakefield GH. 1997. Electrode discrimination and speech recognition in postlingually deafened adult cochlear implant subjects. The Journal of the Acoustical Society of America 102(6):3673–3685

[52] Pfingst BE, Xu L, Thompson CS. 2004. Across-site threshold variation in cochlear implants: Relation to speech recognition. Audiology and Neuro-Otology 9(6):341–352

[53] Noble JH, Gifford RH, Hedley-Williams AJ, Dawant BM, Labadie RF. 2014. Clinical evaluation of an image-guided cochlear implant programming strategy. Audiology and Neurotology 19(6):400–411

[54] Schvartz-Leyzac KC, Zwolan TA, Pfingst BE. 2017. Effects of electrode deactivation on speech recognition in multichannel cochlear implant recipients. Cochlear Implants International 18(6):324–334

[55] Arenberg JG, Parkinson WS, Litvak L, Chen C, Kreft HA, Oxenham AJ. 2018. A dynamically focusing cochlear implant strategy can improve vowel identification in noise. Ear and Hearing 39(6):1136–1145

